# COVID-19 vaccine hesitancy among undergraduate medical students: results from a nationwide survey in India

**DOI:** 10.1101/2021.03.12.21253444

**Authors:** Jyoti Jain, Suman Saurabh, Akhil Dhanesh Goel, Manoj Kumar Gupta, Pankaj Bhardwaj, Pankaja Ravi Raghav

## Abstract

COVID-19 vaccine was launched in India on 16 January 2021, prioritizing health care workers which included medical students. We aimed to assess vaccine hesitancy and factors related to it among undergraduate medical students in India. An online questionnaire was filled by 1068 medical students across 22 states and union territories of India from 2 February – 7 March 2021. Vaccine hesitancy was found among 10.6%. Concern regarding vaccine safety and efficacy, hurried testing of vaccines prior to launch and lack of trust in government agencies predicted COVID-19 vaccine hesitancy. Risk perception regarding contracting COVID-19 vaccine reduced COVID-19 vaccine hesitancy as well as hesitation in participating in COVID-19 vaccine trials. Choosing between the two available vaccines (Covishield and Covaxin) was considered important by medical students both for themselves and their future patients. Covishield was preferred to Covaxin by students. Majority of those willing to take the COVID-19 vaccine felt that it was important for them to resume their clinical posting, face-to-face classes and get their personal life back on track. Around three-fourths medical students viewed that COVID-19 vaccine should be made mandatory for both health care workers and international travellers. Prior adult vaccination didn’t have an effect upon COVID-19 vaccine hesitancy. Targeted awareness campaigns, regulatory oversight of vaccine trials and public release of safety and efficacy data and trust building activities could further reduce COVID-19 vaccine hesitancy among medical students.

## INTRODUCTION

COVID-19 has emerged as a global pandemic with 113 million confirmed cases and 2.5 million deaths worldwide, as on 2 March 2021.^1^ As a part of control measures against COVID-19, vaccines have been launched in India from 16 January 2021.^2^

In the first phase, health care workers including medical students are targeted for vaccination with either of the two vaccines approved for restricted emergency use - Covishield or Covaxin. Covishield is manufactured by Serum Institute of India under license from Astra Zeneca (adenovirus vectored ChAdOx1 nCoV-19 vaccine - AZD1222)^3^ whereas the inactivated SARS-CoV-2 vaccine Covaxin (BBV152) is manufactured in India by Bharat Biotech in collaboration with Indian Council of Medical Research.^4^ Subsequently, from March 1 2021, COVID-19 vaccination has been extended to those aged more than 60 years and those with comorbidities from 45-59 years of age.^2^ The process of registration for the vaccination is done online through the COVID-19 Vaccine Intelligence Network (CO-WIN) portal which is developed with the support of UNDP.^5^ It is also configured to track enlisted beneficiaries, issue SMS reminders and vaccination certificates for users.^5^

Vaccine hesitancy has been frequently studied among health care workers and especially medical students.^6^ The COVID-19 pandemic spurred the rapid development of COVID-19 vaccines with their prominent coverage in news and social media.^7^ Recent studies highlighted the concerns regarding adverse events, unduly rapid vaccine development and poor vaccine efficacy as some of the possible reasons for vaccine hesitancy among medical students.^8–12^ In the Indian situation, out of the two vaccines, the safety and phase 3 efficacy data was publicly released only for Covishield through a scientific publication of the parent Astra Zeneca vaccine.^13,14^ For Covaxin, only the safety and immunogenicity data of phase 1 trial is available.^15^ An announcement of 81% efficacy has only been recently made on 3 March 2021^4^, while its scientific publication is awaited. Although provided free of charge, there has been no option for the health workers to choose between the two vaccines since allocation of vaccines to health facilities had been centrally determined owing to limited supply. Therefore, considering the rapidly evolving situation, the study of vaccine hesitancy among medical students is important. The present study aims to assess the awareness and sources of vaccine information, attitudes and possible determinants of COVID-19 vaccine hesitancy among medical students enrolled in MBBS course in India.

## METHODS

A cross-sectional study was conducted among the cohort of undergraduate medical students in India for a period of around 5 weeks from 2 February – 7 March 2021.

Sample size was calculated pertaining to the prevalence of COVID-19 vaccine hesitancy or refusal among medical or nursing students from previous reports which ranged from 6% in Egypt, 13.9% in Italy, 23% in USA, 30.5 in Malta.^9,16–18^ This yielded a sample size of 962 individuals corresponding to the lowest prevalence, relative precision of 25% and alpha error of 5%.

An anonymous online structured questionnaire was prepared using evidence from prior studies on vaccine hesitancy in general^19,20^ and COVID-19 vaccine hesitancy in medical students.^11^ The questionnaire consisted of three main sections – first section with basic demographic details and assessment of awareness and source of information regarding COVID-19 vaccine, second section with assessment of attitudes regarding the vaccine and the third section relating to prior vaccination experience. This questionnaire was deployed online using google forms. The link for the survey form was exclusively shared with the social network of undergraduate medical students – both individually and through their social media groups. Non-probability sampling strategy was used to target all medical students consenting and willing to spare the time to fill the survey.

Upon completion of the survey, data was downloaded in comma-separated values format and data analysis was conducted using SPSS software version 23.0. Categorical variables related to the survey items were tabulated and odds ratio for vaccine hesitancy was calculated using univariate approach. Subsequently, multivariate logistic regression was conducted to test for plausible determinants of vaccine hesitancy while adjusting for gender, type of medical college, being in pre-clinical or clinical part of course and lack of prior vaccine experience. Similar analysis was repeated for exploring the determinants of hesitancy of joining COVID-19 vaccine trial. A p-value of less than 0.05 was taken as significant. Data analysis was done using STATA v11 and EpiInfo™ v7.2.4.

The study has been approved by the Institutional Ethics Committee of All India Institute of Medical Sciences (AIIMS) - Jodhpur, India (Ref: AIIMS/IEC/2021/3438). Data collection was completely anonymous with no individual level information or name of medical college being collected.

## Results

A total of 1068 students from 22 states and union territories of India participated in the online survey (Fig 1). Around four-fifths of students were from Rajasthan state (Table 1). Gender of students were almost equally distributed (48.6% females). Nearly one-fourths of students were studying in the clinical part of the MBBS course (Table 1).

**Table 1:**
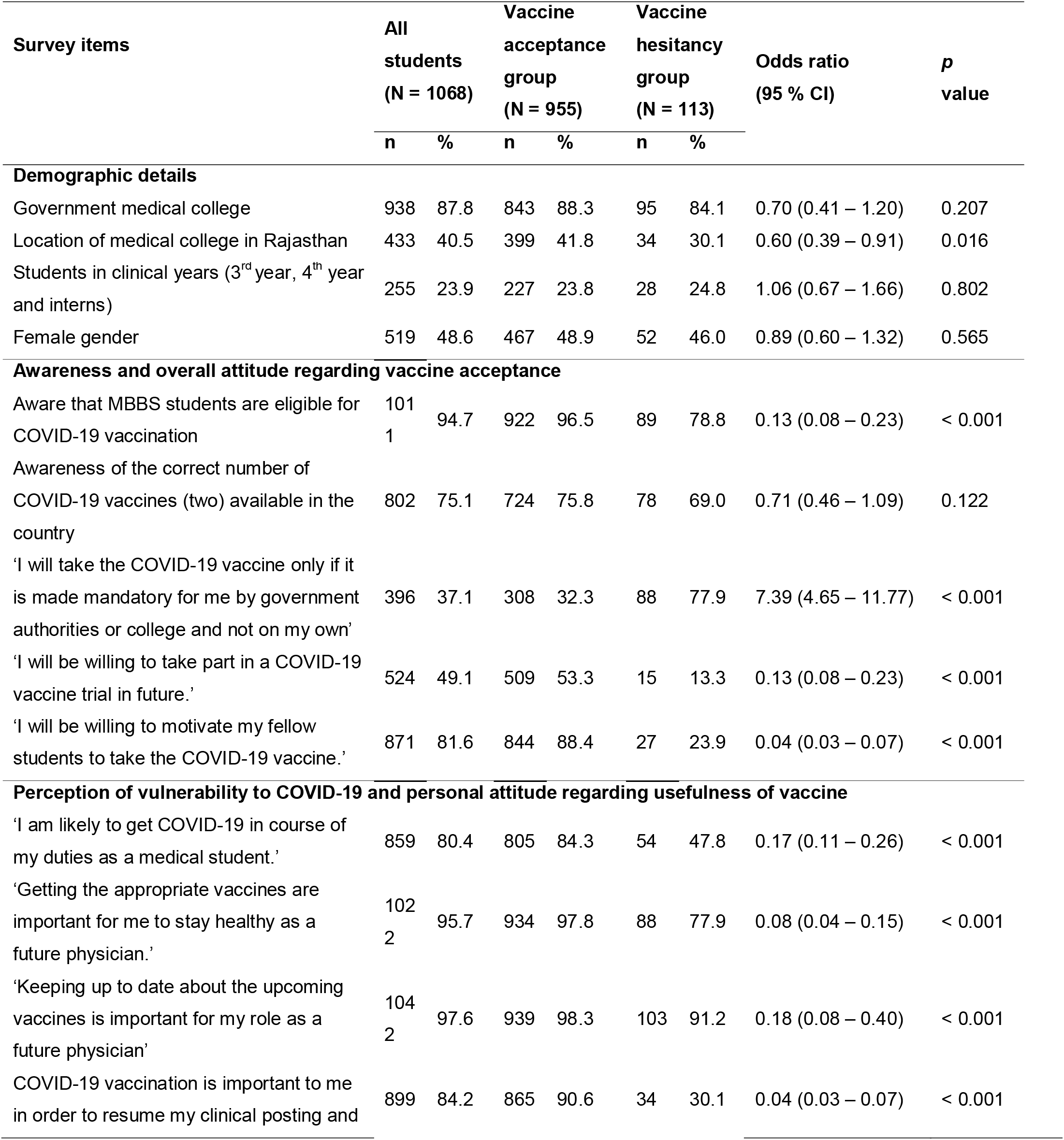
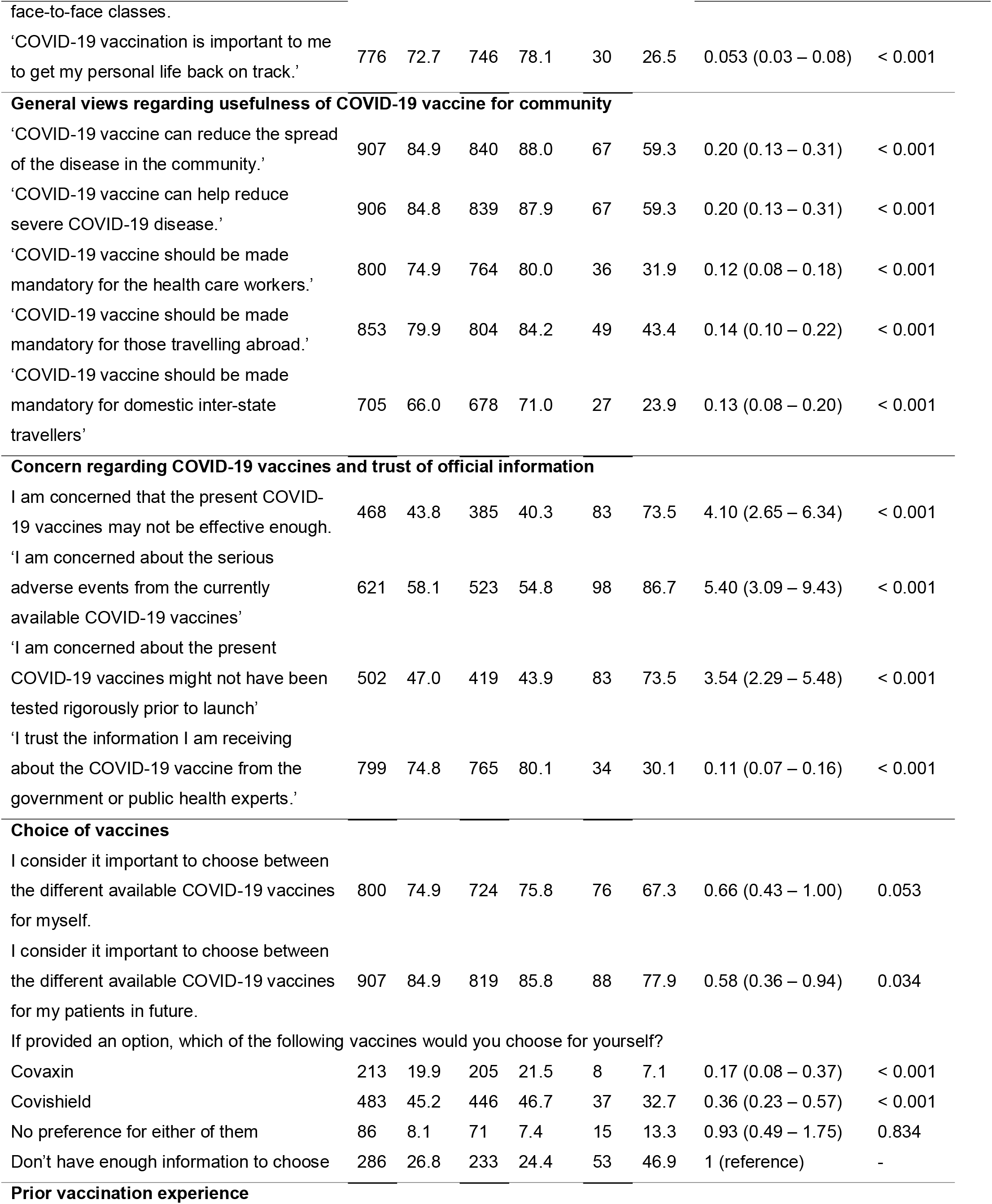
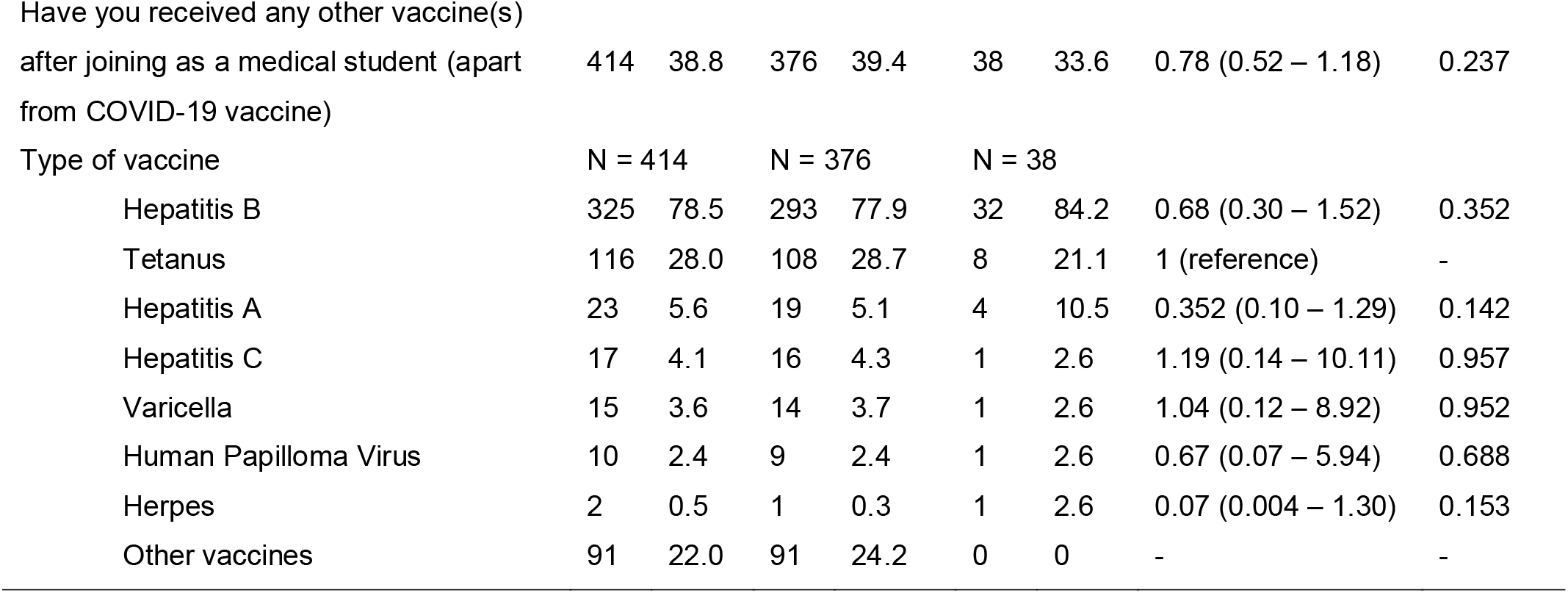
Responses of medical students belonging to vaccine acceptance and hesitance groups (N = 1068)

**Figure 1:**
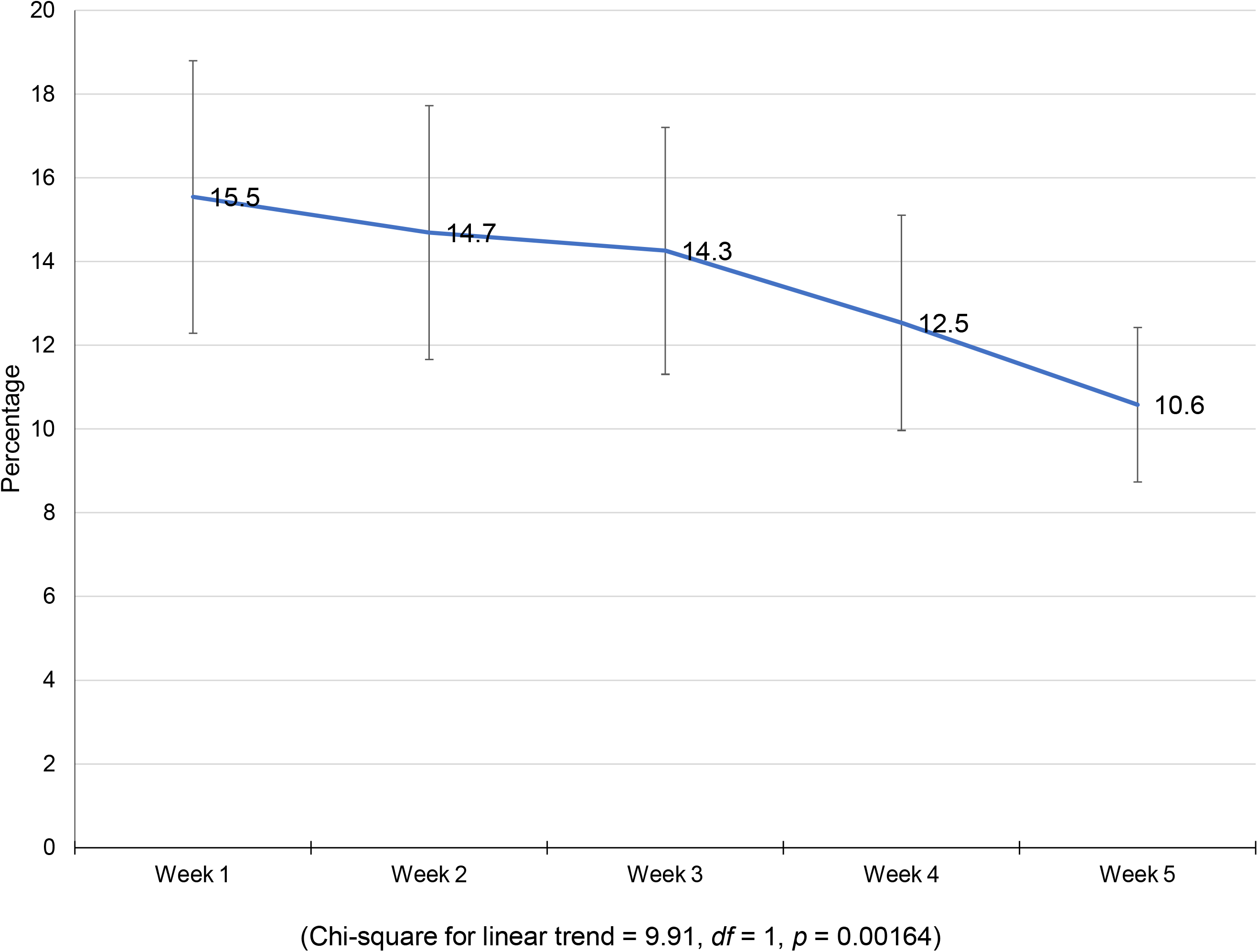
State/Union territory-wise participation of medical students in the COVID-19 vaccine survey (n = 1068)

In response to the statement ‘I am willing to take the COVID-19 vaccine when offered’, 43 (4.0%) ‘disagreed’ and 70 (6.6%) were ‘not sure’. Therefore, vaccine hesitancy was found among 113 students (10.6%). Among those who agreed, 689 (64.5%) had already taken the vaccine and 266 (24.9%) were yet to receive the vaccine at the time of responding to the survey. Cumulative vaccine hesitancy based on the online responses showed a significant declining trend (*p* = 0.00164) from 15.5% at the end of the first week of the survey to 10.6% at the end of the fifth week (Fig 2). Internet, social media and teachers at medical college were the most common source of information regarding COVID-19 vaccine for both the vaccine hesitance and acceptance groups (Fig 3). Further, we found no significant difference between the sources of vaccine-related information between the vaccine acceptance and hesitance groups (Fig 3). Concern regarding safety of COVID-19 vaccine followed by concern regarding its efficacy was the most common reason cited by those hesitant to take the vaccine (Fig 4).

**Figure 2:**
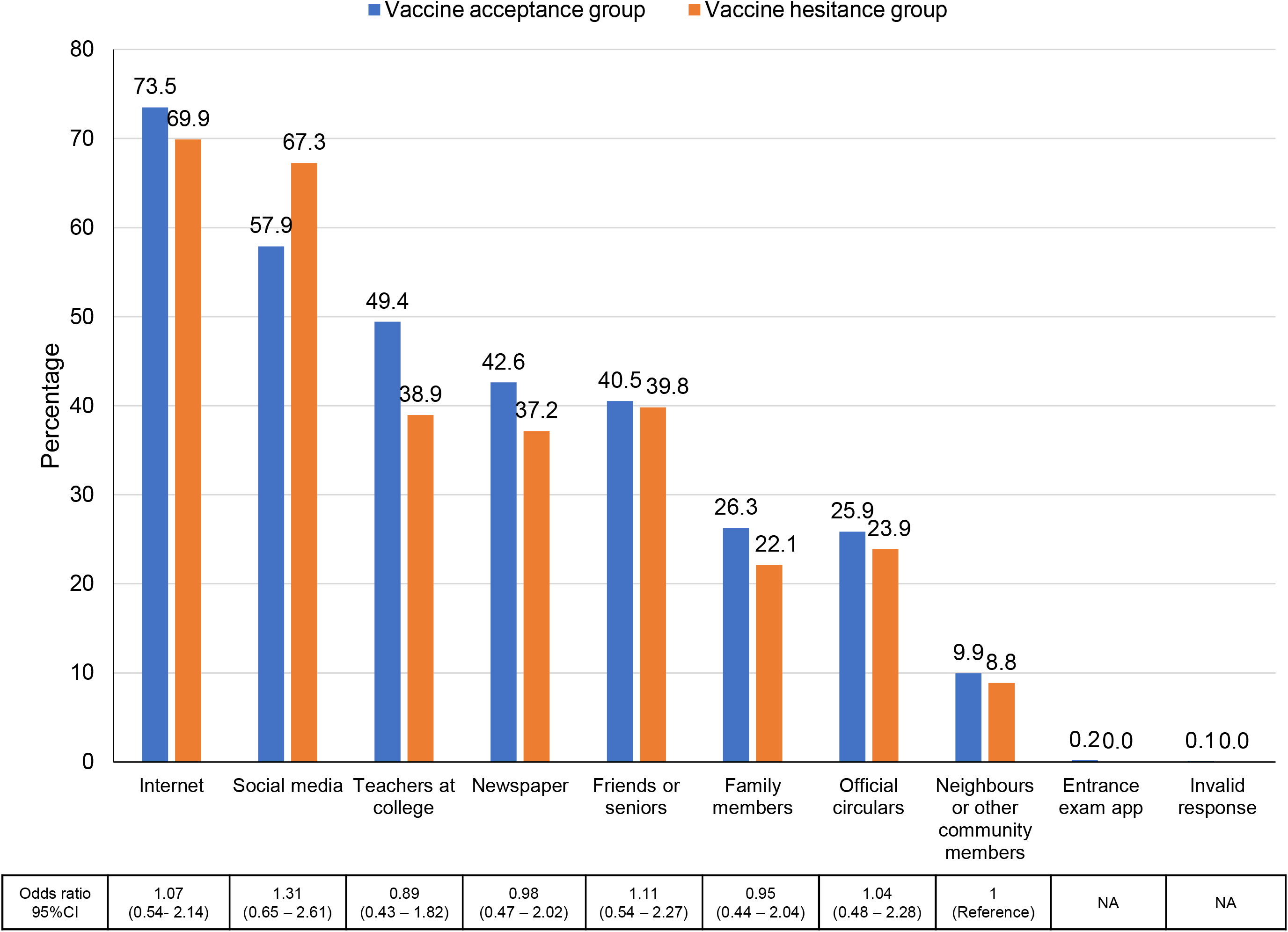
Week-wise trend of cumulative COVID-19 vaccine hesitancy among surveyed medical students

**Figure 3:**
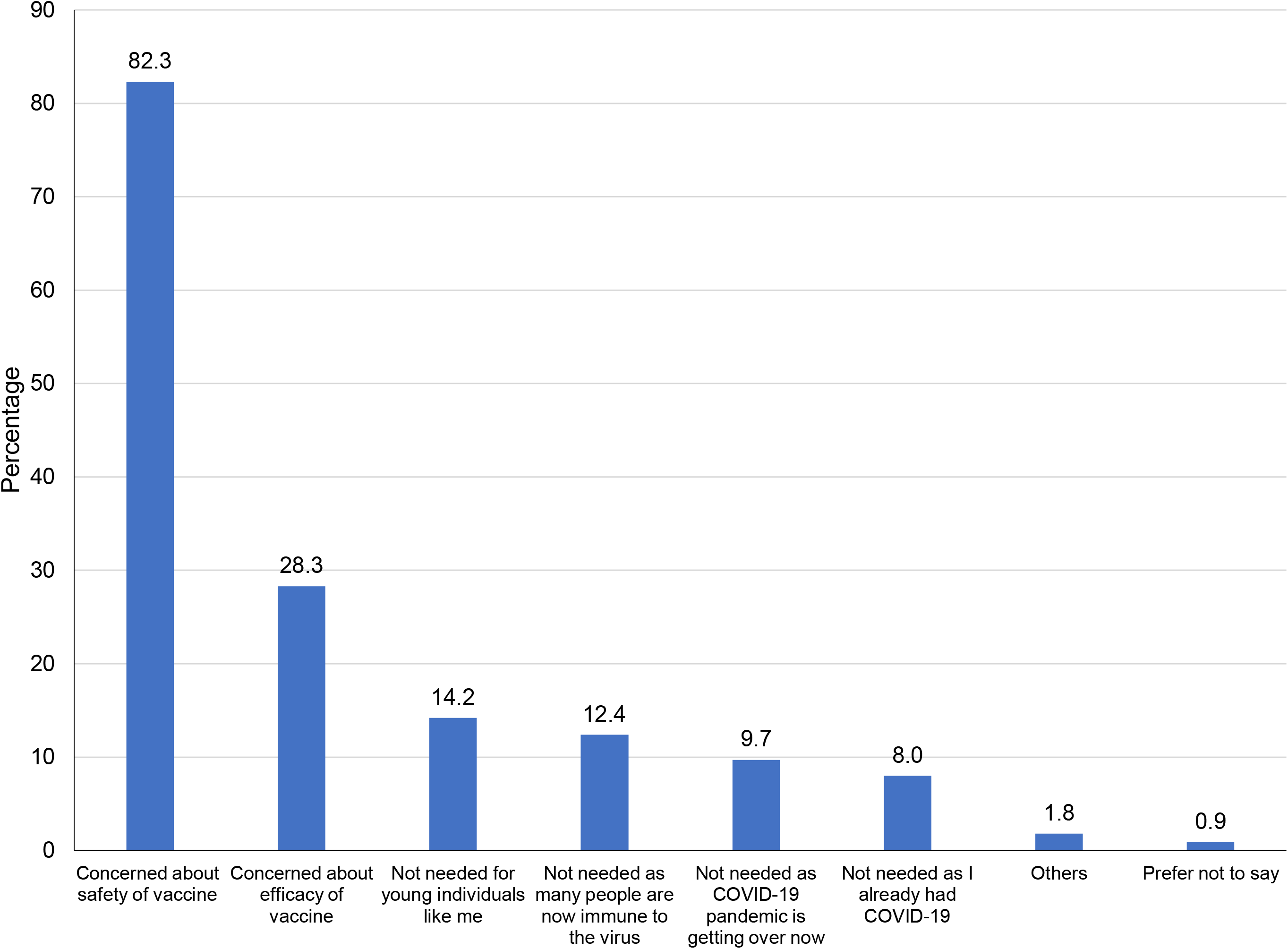
Sources of information regarding COVID-19 vaccine for the medical students (n = 1068)

**Figure 4:**
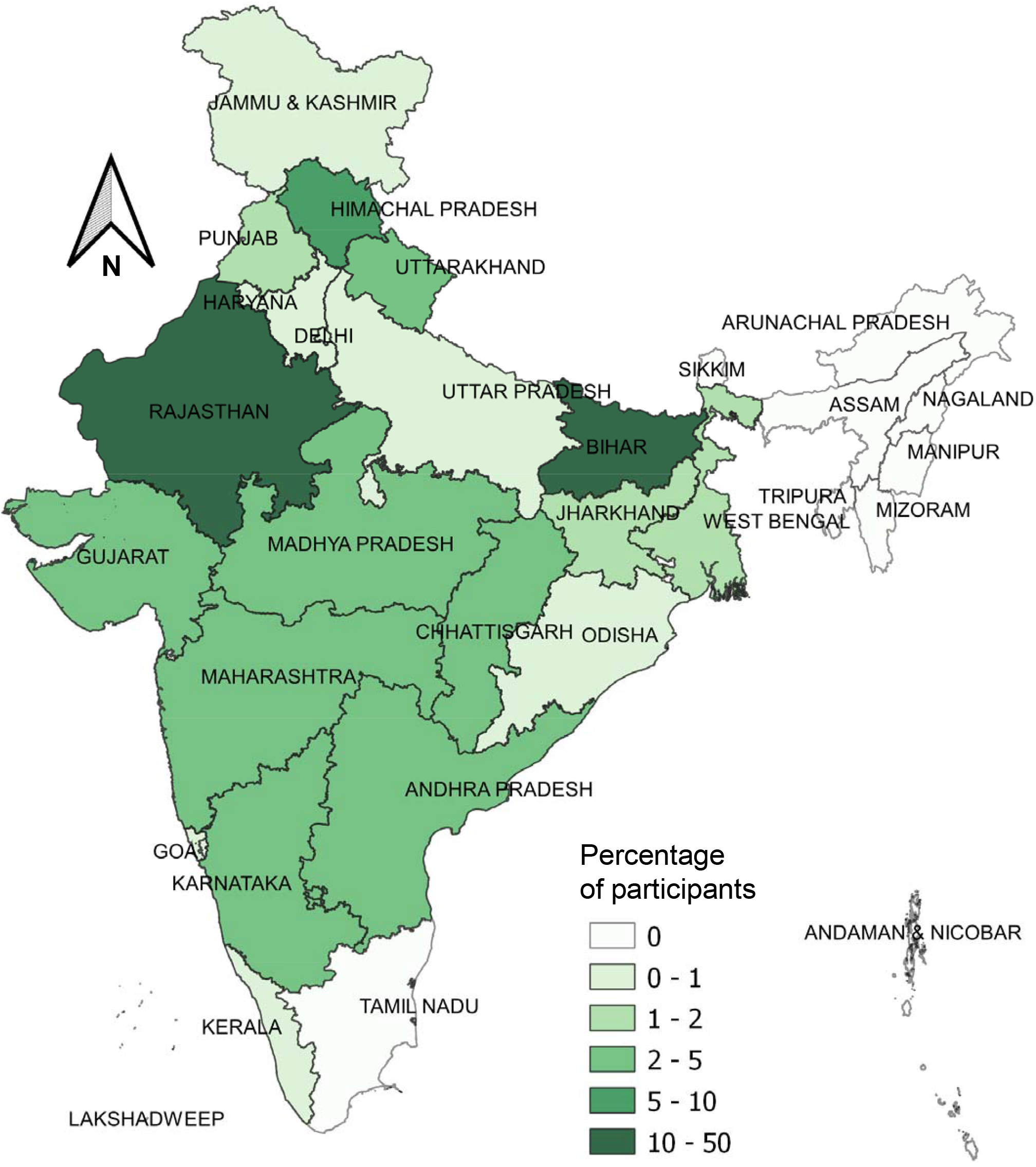
Reasons for COVID-19 vaccine hesitancy among the medical students (n = 113)

Upon conducting logistic regression, lack of awareness of medical students regarding their COVID-19 vaccine eligibility, concern regarding vaccine safety and efficacy and lack of trust in public health authorities were associated with COVID-19 vaccine hesitancy (Table 2). Hesitation in joining COVID-19 vaccine trial was predicted by lack of trust in government or public health authorities (Table 3). Conversely, presence of risk perception among students regarding COVID-19 was associated with lesser hesitancy in taking COVID-19 vaccine as well as joining COVID-19 vaccine trials (Table 2, Table 3).

**Table 2:** Multivariable logistic regression for plausible determinants of COVID-19 vaccine hesitancy (N = 1068)

| Variables | Odds ratio (95% CI) | p value |
| --- | --- | --- |
| Studying in government medical college | 1.32 (0.82 – 2.13) | 0.243 |
| Studying in clinical year | 0.99 (0.57 – 1.72) | 0.966 |
| Male gender | 1.33 (0.82 – 2.13) | 0.244 |
| Lack of awareness regarding eligibility of medical students for<br>COVID-19 vaccination | 4.08 (1.97 – 8.45) | < 0.001 |
| Presence of risk perception regarding COVID-19 | 0.18 (0.11 – 0.30) | < 0.001 |
| Prior vaccination experience present | 0.97 (0.59 – 1.60) | 0.908 |
| Concern regarding adverse effect of vaccine | 3.63 (1.86 – 7.07) | < 0.001 |
| Concern regarding efficacy of vaccine | 2.23 (1.30 – 3.84) | 0.004 |
| Lack of trust in govt. or public health authorities | 5.93 (3.68 – 9.56) | < 0.001 |

Model parameters: Log likelihood = - 247.59, Minus 2 log likelihood difference vs. intercept = 226.05, df = 9, p < 0.0001. Pseudo R-square = 0.3134

**Table 3:** Multivariable logistic regression for plausible determinants of hesitancy regarding participation in COVID-19 vaccine trials (N = 1068)

| Variables | Odds ratio (95% CI) | p value |
| --- | --- | --- |
| Studying in government medical college | 1.14 (0.77 – 1.70) | 0.513 |
| Studying in clinical year | 1.43 (1.05 – 1.95) | 0.024 |
| Male gender | 0.91 (0.71 – 1.17) | 0.478 |
| Lack of awareness regarding eligibility of medical students for COVID-19 vaccination | 1.45 (0.79 – 2.66) | 0.234 |
| Presence of risk perception regarding COVID-19 | 0.34 (0.24 – 0.49) | < 0.001 |
| Prior vaccination experience present | 1.14 (0.88 – 1.49) | 0.320 |
| Concern regarding adverse effect of vaccine | 0.95 (0.71 – 1.27) | 0.713 |
| Concern regarding efficacy of vaccine | 1.18 (0.88 – 1.58) | 0.264 |
| Lack of trust in govt. or public health authorities | 2.33 (1.71 – 3.17) | < 0.001 |
| Model parameters: Log likelihood = - 691.08, Minus 2 log likelihood difference vs. intercept = 98.03, df = 9, p < 0.0001. Pseudo R-square = 0.0662 |  |  |

Comments by medical students were arranged in four themes – ‘confidence in vaccine’, ‘concern regarding vaccine’, ‘practical considerations’ and ‘need for better education’ (Table 4).

**Table 4:** Comments provided by medical students regarding COVID-19 vaccine.

| Theme (total number of comments) | Representative Quotes |
| --- | --- |
| **Concerns regarding vaccine** | 'Adverse effects of vaccine are too much.'<br>'We still don't know the long-term effect of this vaccine so the people who are in low covid prone area can avoid taking the vaccine until clinical trial ends & rest who are at high risk of exposure to COVID-19 should take the vaccine. In this way it won't affect the entire population'<br>'Vaccine is being administered without consent especially to the medical students, which is not ethically correct, given the unavailability of adequate data regarding its safety. My registration was done by the college authorities without my prior knowledge and without asking about my medical background e.g., if I |
|  | have any coexisting diseases which could probably be related to increased risk of any adverse effects of the vaccine. Nor were we warned about the expected side effects like fever and injection site pain.'<br>'I just wanted that Govt of India makes sure that these vaccines have gone through complete and proper testing/trial before they start being used widely.'<br>'It's efficacy and side effects should be mentioned properly, updated information.'<br>'Heard, Covishield is fully tested vaccine till last phase and is a formula from Oxford, but I'm concerned about Covaxin, which they didn't complete the last phase of trial (rumour) and is manufactured in India and is being given.'<br>'It should not be mandatory for any one'<br>'I am worried about the side effects of the vaccine'<br>'I doubt Its potential'<br>'There are some questions which can be answered rather than this or that. I mean to express our own thoughts and.. I am bit concerned of the vaccines as they are not properly tested under the trials but I don't mind if it causes minute problems. And I am ready for the trials if wanted.'<br>'The socioethical condition, family condition of student must be considered and Side effects must be taken into account.'<br>'It would be very good if this vaccination trail is transparent and the data and efficacy should be approved by WHO and then given trials on people...hoping for a better tomorrow with COVID-19 free world'<br>'Govt and medical officials failed to provide proper info to common folks about mechanism of action of vaccines.'<br>'I hope it works.' |
| **Confidence in vaccine** | 'I think to control the spread of this disease, vaccination is must and everyone should participate in it'<br>'Vaccine should be made available for the general public as soon as possible.'<br>'Vaccination is must. Only believe the experts and not the fake media or social influencers. Side effects are just as same as other vaccination like fever, body pain, etc which is quite normal. Be mentally strong before vaccination as being mentally strong is very important for your health. And being a medical student, it is your duty to get vaccinated.'<br>'Just go for it'<br>'We as health care professionals should avoid rumours about COVID-19 vaccine as they don't have any proper base and should believe in our researchers, doctors and government' |
|  | 'It's awesome, thanks for the adventure'<br>'Do it as fast as possible'<br>'We medical students should encourage everyone to being vaccinated'<br>'Vaccination is important to all the country.'<br>'Proud to say that it's our own.' |
| **Practical considerations regarding vaccination** | 'Those below 20 should not be given vaccine according to me!!'<br>'Don't be vaccinated during exam because it may cause variable side effect'<br>'There should be certain blood parameters to determine if the vaccine has generated certain immune response or not'<br>'I think it would be better to have a test done for each one regarding the vaccine. Because the vaccine can act differently in different persons. And if he has already immunised himself by his body defence system. I think no need to take vaccine then... '<br>'Our vaccination is yet pending'<br>'Colleges should take better part in organising vaccines, and assigning authority for management and information on where to receive vaccines as many days vaccination site is not created in college itself, so management should take into consideration students' comfort as classes are already without significant social distancing'<br>'Because of Covid Vaccine our exams are being started in hurry. So much less preparation' |
| **Need for better education regarding vaccine** | 'The medical institutions should inform about the dos and don'ts after putting vaccine.'<br>'All the effects after virus should be informed. So that people don't consider them as side effects and advise against it'<br>'I think data analysis should be made clearer. Or it might be possible that I am not going to the right articles?'<br>'Which vaccine is best right now?'<br>'Because of less awareness about vaccination, number of vaccinated people are less so we should create awareness about COVID-19 vaccination, its benefits and its effect on our country.'<br>'Please make this type of forms for spreading awareness of different types of vaccines, basic differences, plus points, minus points.' |

## Discussion

### Awareness of COVID-19 vaccine and sources of information

COVID-19 vaccination provides a renewed opportunity to closely study the dynamics of health behaviour change in a well-informed young adult population. We found that better awareness regarding the COVID-19 vaccine was associated with reduced hesitancy, similar to study conducted earlier.^6^ It is important to note that vaccine hesitancy for newly launched vaccines reduced over time in our study, which has also been observed earlier.^21,22^ COVID-19 vaccine uptake, especially among young college students has been explained through diffusion of innovation theory through openness to experience and adoption of descriptive norm.^21^ Innovators and early adopters of COVID-19 vaccination could play a role in facilitating its wider acceptance in the medical student community.^23^

The views of students should also be seen in the matrix of multiple sources of information available to them. Our findings support that the role of internet and social media as an information source of health behaviours has been increasingly important for medical students.^19^ Any future intervention to reduce vaccine hesitancy among the student population should take into account this realignment of sources of information. Since the sources of vaccine information were not different among the vaccine acceptance and hesitance groups, we don’t recommend promoting or restricting any particular information channel to tackle vaccine hesitance among medical students.

### Determinants of vaccine hesitancy

Adoption of vaccination practices by healthcare workers plays a key role in motivating the general population through setting of example.^8,24^ Concerns regarding COVID-19 vaccine adverse events as a possible reason for hesitancy has been highlighted by most studies concerning both university students and general population.^8–12,17^ Further, concerns regarding vaccine efficacy seen to play a role in adoption of the COVID-19 vaccine.^8,10,17^ The real concern regarding adverse events appeared to be from the possible ‘long term’ effect of the vaccine. This was coupled with the apprehension that the vaccines had not been tested rigorously enough to determine all possible adverse events and efficacy in a proper manner. The short-term adverse events were also inconveniencing the students owing to vaccination sessions held close to their examinations. The concern regarding vaccine adverse events and efficacy were further elaborated by the comments provided by students. Additionally, concern of lack of consent for provision of data for registration of COVID-19 vaccine by medical students was also observed.

Overall, more than three-fourths medical students viewed that COVID-19 vaccine should be made mandatory for both health care workers and international travellers. However, those hesitating to take COVID-19 vaccination were also less convinced about the various aspects of usefulness of the vaccine for the community such as its potential in reducing the spread of infection or severe COVID-19 disease. They were also much less likely to have it mandated for health care workers and domestic and international travellers.

Majority of even those hesitating displayed a sense of responsibility in their role as future physicians to keep up to date regarding the upcoming vaccines and their importance to keep themselves healthy. This suggests that hesitation regarding COVID-19 vaccination could be related to issues specific to it rather than due to apathy towards vaccines in general. Therefore, targeted education and trust building by regulatory agencies and medical colleges could help reduce COVID-19 vaccine hesitancy considerably.

Our findings also seemed to match with the health belief model^23^ wherein the perceived susceptibility to COVID-19 and perceived benefits of vaccination had a role in lessening the hesitancy for COVID-19 vaccination. We also found that a sizeable proportion of students had indeed received the vaccination despite having concerns which indicated that acceptance of vaccination was not purely voluntary. It appears unlikely that this coercion could be entirely driven by the pressure of college authorities. Within this framework, COVID-19 vaccine acceptance could have been a subjective norm and pressure of social conformity could have influenced some hesitant students to finally get vaccinated. Further, majority of those choosing to be vaccinated were motivated by desire for resumption of clinical and face-to-face classes by the prospect of getting their personal lives back on track. Therefore, COVID-19 vaccination was also seen a confidence building measure which could help the students ease their restricted life during COVID-19 pandemic. Confidence regarding the vaccine was also expressed by the students as free comments. On the other hand, those hesitating were much less likely to believe in this enabling effect of COVID-19 vaccination.

Concern for adverse events didn’t deter medial students to participate in vaccine trials unlike their counterparts in the United States of America.^11^ Risk perception of self-regarding COVID-19 increased the students’ willingness to participate in COVID-19 vaccine trial. On the other hand, lack of trust in government or public health authorities deterred them from participating in vaccine trials, similar to what was observed in previous studies. ^11,25,26^

### Choice of vaccines and previous vaccination

Students considered it important to choose between the available COVID-19 vaccines both for themselves and for their future patients. Between the two available vaccines, Covishield was preferred whereas a considerable proportion also felt that they didn’t have enough information to choose. Acceptance of Covaxin was found to be less in general and was even lesser among those hesitating to take the vaccine. This situation might change in future with more information on safety and efficacy vaccines being available.

Experience of prior vaccination has been found to have a role in increasing the acceptance of COVID-19 vaccine.^9,25,27^ However, this was not replicated in the present study. This could be mainly because in the present setting, Hepatitis-B was the vaccine taken by majority of the students unlike the studies from outside India in which annual Influenza vaccination had been considered.^9,25,27^ Since the importance of Hepatitis B vaccine is well-accepted for healthcare professionals, its uptake might be more related to medical colleges’ policy of offering vaccination to medical students during their course rather than vaccine hesitancy *per se*.

### Implication of findings for general population

Care needs to be taken while extrapolating the findings among health care workers and medical and nursing students to the general population. This is since acceptance of vaccine might be more among medical students as compared to non-medical students and general population.^24^ It was also found that conspiracy theories don’t tend to affect medical students as compared to non-medical students.^24^ On the other hand, a study conducted in Italy found no difference between hesitancy among medical and non-medical studnets.^18^ Therefore, transferability of findings among medical students to the community appears to be context-specific. Challenges faced in the community may be more regarding provision of accurate information and tackling vaccine hesitancy in general whereas among health workers and medical students it would be mainly related to safety and efficacy issues specific to the newly launched vaccines.

### Limitations

Our survey had the limitation that it was conducted after COVID-19 vaccination had started in some of the medical colleges. Therefore, it could have underestimated the initial vaccine hesitancy for those who were vaccinated and would have subsequently converted to the vaccine acceptance group. Although we captured students’ responses through open comments, the online mode of data collection often fails to capture the depth of information which could have otherwise been possible through qualitative methods applied in face-to-face settings.

## Conclusions

COVID-19 vaccine hesitancy was found in one out of every ten medical students. Lack of awareness regarding vaccination eligibility, concern regarding adverse events and efficacy of the vaccine and lack of trust in government were independently predictive of vaccine hesitancy. Heightened risk perception regarding COVID-19 reduced vaccine hesitancy.

Concerns regarding lack of vaccine-related information and launch of vaccine prior to release of safety and efficacy data were noted. Although vaccine hesitancy showed a diminishing trend over time, health education programmes tailored to boost awareness regarding vaccine and improve trust in government agencies would be helpful. Taking due informed consent for registration of personal information in vaccine portal and ensuring that vaccination sessions are not held just before examinations could further improve acceptance of newly launched vaccines. As future health care providers, concerns of medical students should be addressed on priority basis.

## Data Availability

Data upon which the study findings are based will be included in Supplementary file 1, along with the journal article upon final acceptance for publication.

## DECLARATIONS

## Authors’ contributions

JJ conceived the idea of the study. JJ and SS designed the data collection format with inputs from MKG, PB. JJ conducted data collection. SS wrote the manuscript with inputs from JJ, MKG, PB, AG and PRR. All authors approved the final manuscript.

## Acknowledgement

We acknowledge the help of medical students who participated in the study.

## Funding

The authors declare that no funding was received from any source for the study and preparation of this article.

## Conflict of interest

The authors declare that there are no conflicts interests for publication of this article. The views expressed in this article are those of the authors alone and do not necessarily represent the views of their organizations.

## Ethical approval

The study has been approved by the Institutional Ethics Committee of All India Institute of Medical Sciences (AIIMS) – Jodhpur, India (Ref: AIIMS/IEC/2021/3438).

## Data availability statement

Data upon which the study findings are based will be included in Supplementary file 1, along with the article upon acceptance for publication.

## Supplementary file

**Supplementary file 1:** Data for ‘Determinants of COVID-19 vaccine hesitancy among undergraduate medical students: results from a nationwide survey in India’

## Notes

### Competing Interest Statement

The authors have declared no competing interest.

### Author Declarations

The study has been approved by the Institutional Ethics Committee of All India Institute of Medical Sciences (AIIMS) Jodhpur, India (Ref: AIIMS/IEC/2021/3438).

